# Partial recovery of amblyopia following fellow eye ischemic optic neuropathy

**DOI:** 10.1101/2021.12.16.21267939

**Authors:** Hannah H. Resnick, Mark F. Bear, Eric D. Gaier

## Abstract

**Background:** Recovery from amblyopia in adulthood following fellow eye (FE) vision loss is a well-known phenomenon. Incidence of recovery varies widely following different FE pathologies and rate of recovery following FE ischemic optic neuropathy (ION) has not been examined. We aimed to determine frequency and degree of improvement in amblyopic eye (AE) visual function following ION in the FE.

**Methods:** We performed a retrospective chart review of patients between 2007-2021 confirmed to have amblyopia and ischemic optic neuropathy in different eyes. Patients with unstable ocular pathology potentially limiting vision were excluded. We compared best-corrected visual acuity (VA) in each eye before and after FE ION over time. For patients with available data, we examined change in perimetric performance over time.

**Results:** Among the 12 patients who met inclusion criteria (mean age 67±8 years), 9 (75%) improved ≥1 line and 2 (17%) improved ≥3 lines. Median time from ION symptom onset to maximal improvement was 6 months (range: 2-101 months). Reliable perimetric data were available for 6 patients. Mean sensitivity improved in the amblyopic eye for all patients, with a mean improvement of 1.9±1.1 dB. There was no correspondence between foci of ION-related field loss and gains in field sensitivity in the AE.

**Conclusion:** A high proportion of patients with amblyopia and contralateral ION experience improvement in their amblyopic eye. Modest gains in perimetric sensitivity in the AE may accompany FE ION. These findings support the view that residual plasticity in the adult visual cortex can be tapped to support functional improvement in amblyopia.

## Introduction

Amblyopia results from abnormal visual experience during early childhood and is a common cause of visual impairment among children and adults [1, 2]. Treatment for amblyopia has long consisted of occluding the fellow, unaffected eye (FE) before 7 years of age, after which treatment responses are limited [3]. Even when treated within this window, amblyopia often persists into adulthood, with an estimated global prevalence of 3.29% among those older than 20 [1]. The presence of residual amblyopia increases the lifetime risk of significant visual impairment [4].

Clinically significant improvement in amblyopic eye (AE) visual acuity (VA) has been widely reported to occur in adulthood in the setting of FE pathology, including cataracts, ischemia, trauma, and other causes of vision loss [5–10]. Younger age, depth of amblyopia, extent of FE vision loss, and AE fixation pattern have all been reported to be associated with greater gains in AEVA, though inconsistently [5–7], and reports of the incidence of this phenomenon vary from 19-90% [5–8].

Anterior ischemic optic neuropathy (ION) is the most common cause of acute optic nerve-related visual loss in adults [11, 12]. To date, only three cases of AEVA improvement following FE ION have been reported [9], and the frequency of amblyopia recovery following FE ION is unknown. Given the relatively high prevalence of both amblyopia and ION, a better understanding of this phenomenon could inform clinical decision-making, especially as experimental therapies intended to promote visual recovery in the ION-afflicted FE are considered.

To address these knowledge gaps, we conducted a retrospective chart review of patients with amblyopia and FE ION with the primary goal of quantifying AEVA improvement. We examined relationships between clinical factors associated with AE recovery in other contexts and analysed previously unexplored relationships in perimetric performance. We hypothesized that FE ION would lead to improvement in the AEVA and visual field sensitivity, and that perimetric improvement would reflect the location and depth of FE visual field loss.

## Methods

### Patients

This study was approved by the Mass General Brigham Institutional Review Board and adhered to the tenets of the Declaration of Helsinki. We examined medical records for patients ≥18 years of age who were diagnosed with both amblyopia and ION. Cases were collected through medical record billing queries (diagnostic codes H53.0, including all subcodes, and H47.01) for patients registered in the Massachusetts Eye and Ear Infirmary electronic medical record system with diagnosis codes entered between 2007 and 2021. Cases were supplemented with provider (EDG) patient lists. Charts were reviewed manually to determine inclusion. To be included patients had to have notes confirming diagnoses of amblyopia and ION in opposite eyes and VA measurements within one month of FE ION onset. Patients were excluded if the onset of FE ION to the month and year could not be determined. Exclusion criteria also included (1) VA of ≥20/20 in the non-ION eye at the time of diagnosis or at the preceding visit, and (2) a history of bilateral amblyopia.

### Chart Review

Patient charts, including scanned outside ophthalmology notes, were manually reviewed for demographic data (age, sex) and ocular history, including history or presence of strabismus and other ocular conditions at the time of FE ION. Laterality of amblyopia was determined as documented in visit notes (case 7 had inconsistent documentation of AE laterality that was isolated to one provider’s notes with multiple other providers in agreement, so the commonly documented laterality was used). Follow-up length was determined by the last visit with recorded VA measurements or by the onset of a new, vision-limiting FE disorder. Timeframes were recorded to the nearest month if within a few days or as ranges between two months otherwise. Graphing and timeframe calculations were performed by rounding the month up if a range was recorded.

If VA measurements prior to FE ION onset were available, AEVA baseline was determined using the best AEVA in the 5 years preceding ION onset. Otherwise AEVA at the first visit following FE ION served as the baseline. Refractive correction worn at each visit was recorded as documented. When refractive correction was not documented, but correction was used in the VA measurement, the worn refractive correction was inferred based on (1) the most proximally measured worn refraction or (2) the most recently provided manifest refraction. Use of pinhole during VA measurement was noted and used as the best-corrected measurement. Snellen values were converted to LogMAR[13] with additional letters scored as ±0.2 from each line. Counting fingers, hand motions, light perception, and no light perception were designated as 2.1, 2.4, 2.7, and 3.0 LogMAR, respectively.[13]

### Visual Fields

Longitudinal Humphry visual field (HVF; 24-2 and/or 30-2 SITA) performance reports were available for a subset of patients at and/or following presentation for FE ION. Given that fixation instability is a defining feature of amblyopia [14], reliability thresholds for exclusion were set as (1) absence of the physiologic blind spot on the grayscale and (2) false positives or negatives ≥20%. To assess for changes in perimetric sensitivity patterns in the AE over time, raw sensitivity values between the first and last available HVFs were compared at each point. The 2 most nasal points on 24-2 HVFs were excluded to allow for homonymous retinotopic comparisons with the FE. Outermost points on 30-2 HVFs were excluded to allow for comparisons with 24-2 fields over time and blind spots (second-most temporal points above and below the horizontal meridian in 24-2 fields) were excluded. Unweighted mean sensitivity at each timepoint was also calculated using raw sensitivities at each included location.

### Statistical analysis

Statistical analyses were conducted using SPSS (IBM, Armonk, NY). Given the lack of normality among baseline LogMAR values (*p*=0.003, Kolmogorov-Smirnov test), comparisons of eye-specific VAs between baseline and follow up visits were assessed using a two-tailed Wilcoxon signed rank test. Distribution of time to best AEVA and follow-up time were also non-normal (*p*=0.004 and 0.019, respectively; Kolmogorov-Smirnov test), so median and range is reported; otherwise mean (SD) is provided. Associations between magnitude of change in AEVA and dichotomous factors were assessed using two-tailed *t* tests. Spearman correlation testing was used to quantify associations between VA change and continuous factors. In all cases, *p*<0.05 was considered statistically significant.

## Results

### Case characteristics

Of 34 potential cases, 18 cases were excluded because of ION and amblyopia in the same eye, lack of clearly documented ION and/or amblyopia in notes (presumed billing error), or lack of available VA data fitting inclusion criteria. Following initial chart review of the remaining 16 cases, 2 were excluded due to uncertainty regarding the etiology of FE optic neuropathy, and 2 were excluded due to significant potential confounders (FE trauma prior to ION in one case and optic disc edema in the AE at the initial evaluation in the other) (**Figure 1**).

**Figure 1.**
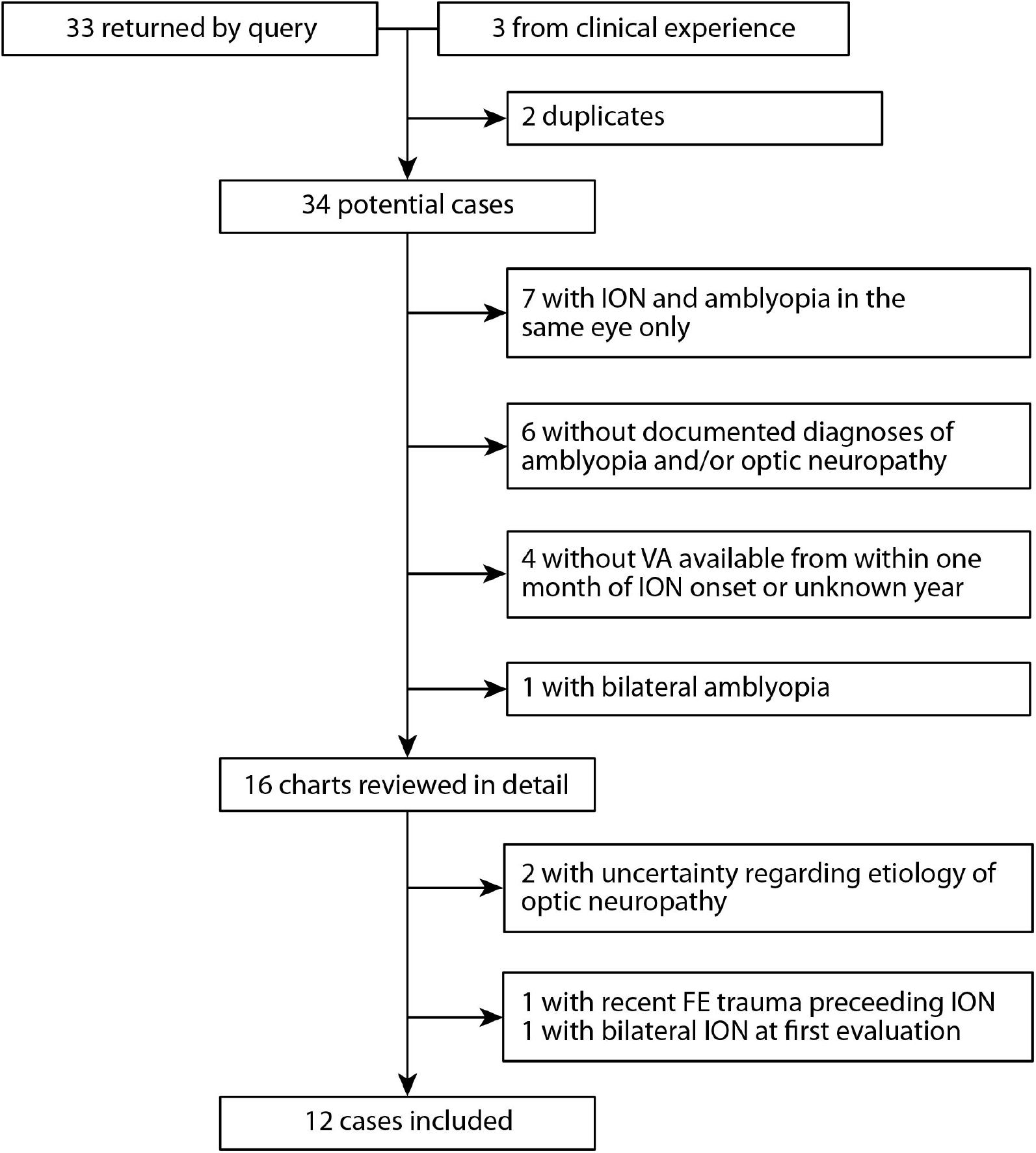
Flowchart depicting patient selection. FE = fellow eye; ION = ischemic optic neuropathy; VA = visual acuity.

Twelve cases were included in the final analysis (**Table 1**). Of these, 2 (cases 3 and 10) had a history of stable ION in the AE (onsets 4 years and within 8 months prior, respectively) and 2 were women (2/12; 17%). Mean (SD) age was 67±8.0. History of occlusion therapy in childhood was noted in 6/12 cases (50%), and 7/12 (58.3%) had current or prior strabismus. ION was treated in 3/12 (25%) patients with systemic corticosteroids. Median length of follow-up was 13.5 months (range: 2-129 months).

**Table 1.** Characteristics and visual acuities of cases of amblyopia with contralateral ION.

| Case # | Age range (years) | Gender | Notable ocular conditions and changes over the course of follow up<sup>a</sup> | Strabismus ever | Systemic Steroids | Baseline AEVA | Best AEVA | AEVA improvement (lines) | Time from ION to best AEVA | Worst FEVA | Follow -up time |
| --- | --- | --- | --- | --- | --- | --- | --- | --- | --- | --- | --- |
| 1 | 70-79 | female | 1. MDF dystrophy, OU, diagnosed after FE ION | yes | yes | 20/600 | 1/20 (20/400) | 1.8 | 4 mo | 20/20- | 4 mo |
| 2 | 70-79 | male | 1. Pseudophakia, OU, prior to FE ION.<br>2. ION, AE, 21 days after FE ION onset | yes | no | 20/150 | no improvement | 0.0 | NA | 20/600 | 12-13 mo |
| 3 | 60-69 | male | 1. Congenital cataracts, OU, not visually significant<br>2. s/p LASIK with mono-vision (AE near, FE far)<br>3. ERM, AE, mild<br>4. ION, AE, ~4 years prior to FE ION | yes | no | 20/40 | 20/25-1 | 1.8 | 2 mo | 20/50-1 | 2-3 mo (3 days after best) |
| 4 | 60-69 | male | 1. Peripheral retinoschisis, FE, prior to ION | no | no | 20/50 | 20/40-1 | 0.8 | 4 mo | 20/250-1 | 11-12 mo |
| 5 | 60-69 | male | 1. Pseudophakia, OU, prior to FE ION<br>2. Posterior capsular opacities, OU, mild<br>3. ERM, OU, mild with lamellar macular hole AE | yes | yes | 20/40 | 20/25-2 | 1.6 | 1 year, 8-9 mo | LP sc | 2 years 8-9 mo |
| 6<sup>b</sup> | 60-69 | female | 1. Pseudophakia, OU, prior to FE ION. | yes | no | 20/30-2 | 20/25- | 1.0 | 8 years, 5 mo | HM | 10 years 8-9 mo |
| 7 | 60-69 | male | None | no | no | 20/25-1 | 20/20 | 1.2 | 2 mo | 20/25-3 | 2 years 1-2 mo |
| 8 | 60-69 | male | 1. Pseudophakia, OU, prior to FE ION. | no | no | 20/40 | No improvement | 0.0 | NA | CF | 2 mo |
| 9 | 40-49 | male | None | no | no | 20/400 | 20/200 | 3.0 | 2-3 mo | 20/50 | 2-3 mo (same as best) |
| 10 | 60-69 | female | 1. ION, AE, timing unknown with pale ON at FE ION onset<br>2. NPDR, OU<br>3. s/p LASIK, OU<br>4. ERM, FE, 4-5 months after FE ION onset (visually insignificant) | yes | no | 20/40 | 20/30-1+2<sup>d</sup> | 1.4 | 8 mo | 20/200 | 1 year 11-12 mo |
| 11 | 70-79 | male | 1. Ptosis, AE | yes | yes | 20/100 | 20/60-2 | 1.8 | 2 years, 7-8 mo | NLP | 2 years 7-8 mo (same as best) |
| 12 | 60-69 | male | None<sup>c</sup> | no | no | 20/50 | 20/20-2 | 3.6 | 9 mo | 20/40 | 4 years 5-6 mo |

<sup>a</sup> Presence of mild and/or overall stable cataracts not included
<sup>b</sup> Inconsistencies in documented laterality of amblyopia
<sup>c</sup> ION with subretinal peripapillary hemorrhage; infectious serologic work-up including Lyme and Bartonella, negative
<sup>d</sup> HOTV linear
AE = amblyopic eye; CF = counts finger; FE = fellow eye; HM = Hand movements; ION = ischemic optic neuropathy; LASIK = laser-assisted in situ keratomileusis; LP= light perception; mo = months; NI = no improvement; NLP = no light perception; NPDR = non-proliferative diabetic retinopathy; PH = pinhole

### Change in AEVA

Median baseline AEVA was 0.35 LogMAR (Snellen 20/40-20/50), range: 0.12-1.48 (20/25-1 to 20/600) (**Table 1**). Baseline AEVA measurements preceded ION onset in 6 cases, of whom 4 had <1 line of difference between the AEVA prior to and at the initial evaluation for FE ION. Baseline AEVA prior to FE ION onset was 2 and 4.9 LogMAR lines greater than that measured at FE ION presentation for cases 3 and 7, respectively, with case 7 using only PH rather than prescribed refractive correction at the time of FE ION presentation.

From baseline through the follow-up period, best AEVA improved significantly by a mean (SD) of 0.15±1.1 LogMAR (1.5 lines, 95% CI: 0.83-2.17 lines, *p*=0.004) (**Figure 2**). Of the 12 cases analyzed, 9 (75%) experienced ≥1 line of AEVA improvement, and 2 (16.7%, cases 9 and 12) exhibited ≥3 lines of improvement. Median time from onset of ION symptoms to best AEVA was 6 months (range: 2-101 months), with 6/9 (66.7%) improved cases reaching their best AEVA within one year.

**Figure 2.**
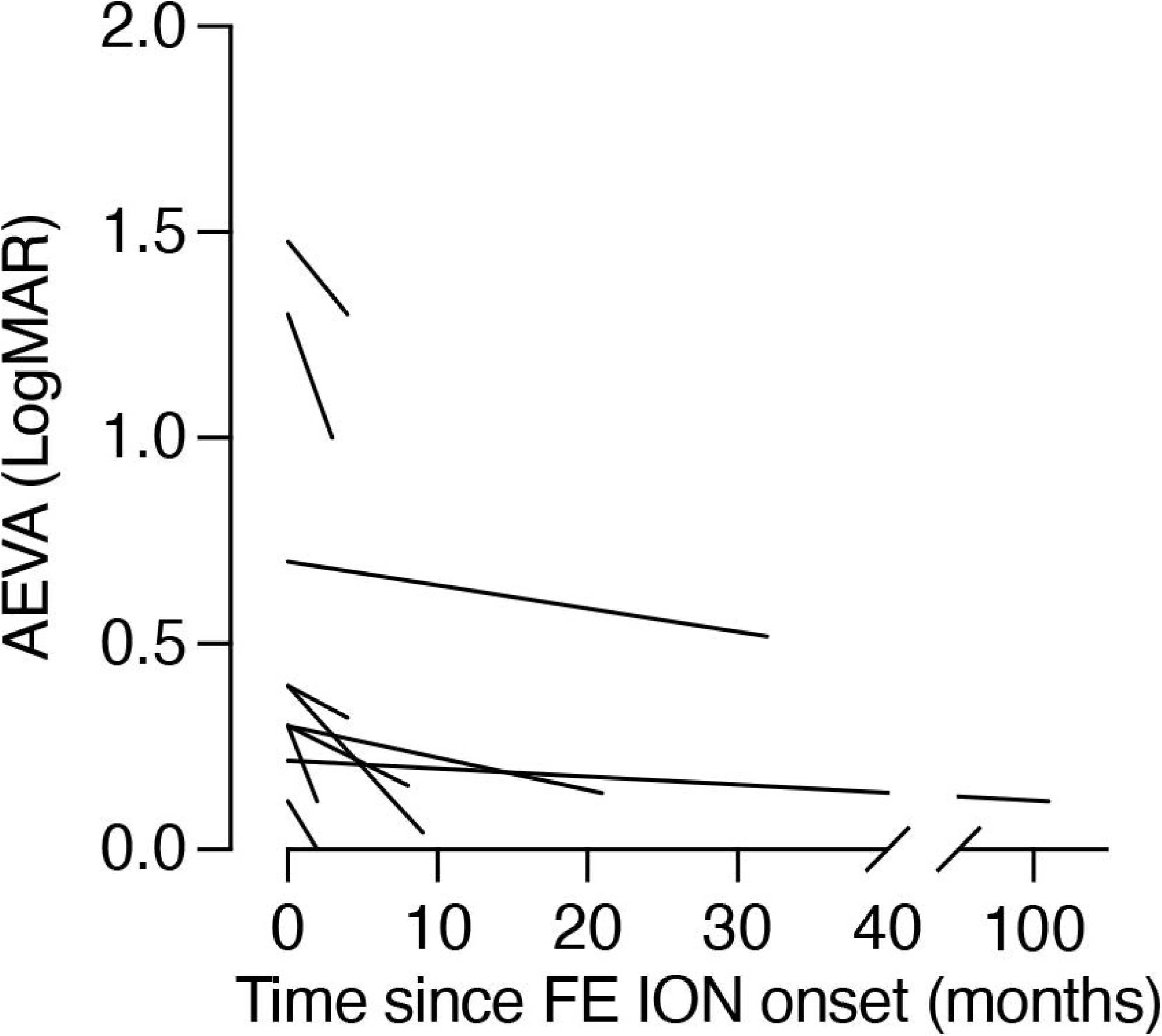
Improvement in amblyopic eye (AE) VA after FE ION. Change in AEVA from baseline to follow up plotted across time since FE ION onset. Values at month 0 represent baselines VA values defined as the best AEVA in the 5 years preceding or at the time of presentation for FE ION.

We next evaluated clinical factors that may contribute to AEVA in the setting of FE ION. Among the 9 patients whose AEVA improved ≥1 line, 8 had available data on their refractive correction at baseline and follow up timepoints. Seven patients used different refractive correction, with or without pinhole, in the AE between baseline and when the best AEVA was recorded; spherical equivalent changed by >1.0 D in 2 cases (1 and 11) (**Supplemental Table 1**). Mean (SD) FEVA nadir was 1.3±1.1 LogMAR. There was no association between baseline AEVA and lines of AEVA improvement (*p*>0.3), nor between FEVA nadir and lines of AEVA improvement (*p*>0.2) (**Supplemental Figure 1)**. Of those with strabismus 6/7 (85.7%) improved ≥1 line in AEVA. There was no difference in magnitude of AEVA improvement between those with strabismus and those without (*p*>0.6). The 3 cases treated with systemic corticosteroids experienced AEVA improvement of 0.16-0.18 LogMAR (1.6-1.8 lines), similar to that seen among those not treated (*p*>0.6).

### Amblyopic Eye Visual fields

Reliable HVFs were available for 6/12 patients (cases 3, 5,7, 10, 11, and 12). Cases 3 and 10 had AE field defects in the setting of prior ION. Mean (SD) unweighted mean sensitivity (MS) at baseline was 24.8±2.9 dB and improved in 6/6 patients by a mean (SD) of 1.9±1.1 dB (**Figure 3**). There was no significant association between change in MS and AE lines of improvement (*p*>0.9). Cases 3 and 11, who exhibited the greatest improvement in MS (+2.24 and +4.04 dB, respectively), both improved 1.8 lines in AEVA. There was no appreciable relationship between the pattern of change in AE HVF sensitivity over time and the acquired visual field deficit in the FE (**Supplemental Figure 2)**.

**Figure 3:**
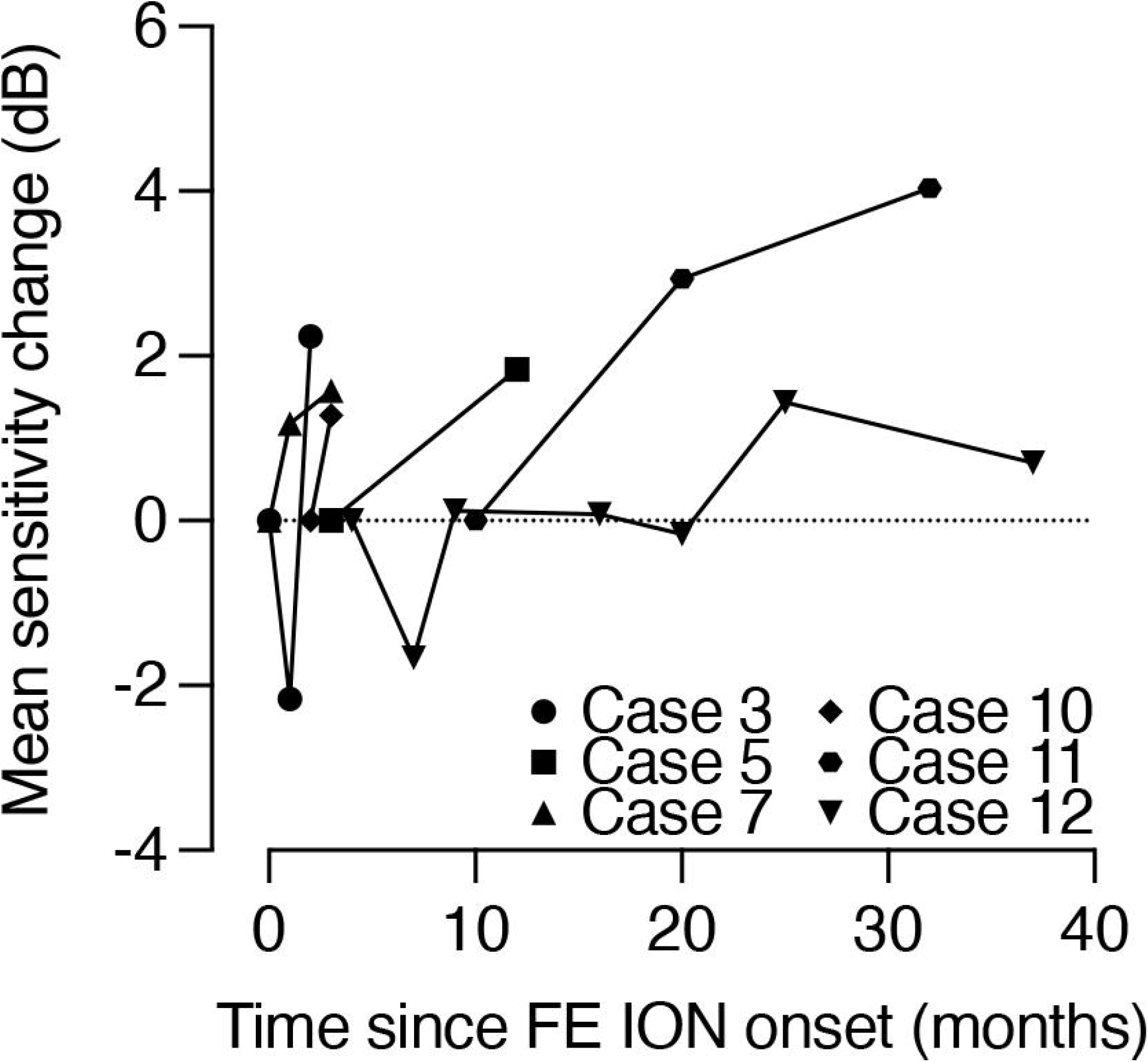
Improvement in AE perimetric sensitivity after FE ION. Change in Humphrey Visual Field (HVF) mean sensitivity in the amblyopic eye from baseline to follow-up plotted across time since FE ION onset for the 6/12 cases with reliable AE HVF data.

## Discussion

Report of patients with amblyopia and FE ION is limited to one published series of 3 cases, all of whom experienced AEVA improvement [9]. Analyzing 12 cases of amblyopia and FE ION, we have significantly expanded the total number of reported cases of this phenomenon and shown that partial but clinically significant AEVA recovery is common.

We found that 75% of patients experienced ≥1 line of AEVA improvement, with 17% experiencing improvement of ≥3 lines. Prior reported incidences of amblyopic eye improvement following FE injury have varied widely. A 1984 European study found an incidence of improvement of 28.5% [5]. In a 2002 United Kingdom study including patients with FE vision loss from multiple causes, Rahi et al. reported only 48/254 (19%) of cases experienced AEVA improvement with 10% improving ≥2 lines [6]. By contrast, studies examining amblyopic eyes in the setting of several specific causes of FE vision loss, rather than any cause of decreased contralateral vision, have found higher rates of recovery: 52% of those with FE uveal melanoma had ≥2 lines AEVA improvement [8], as did 90% of those with age-related macular degeneration [7]. Wide variance in reported AEVA recovery may suggest that certain types of contralateral eye pathologies are stronger drivers of recovery than others.

Closure of the critical period of visual development is defined by an increase in the threshold for the neuroplasticity necessary for recovery from amblyopia [15]. This threshold is apparently lowered by damage to the FE. Ocular pathologies that similarly limit visual function in the FE may involve drastically different effects on retinal ganglion cell function, the visual signals transmitted to the brain (e.g. cataract versus ION), and the potential for recovery from amblyopia. Precisely how different forms of FE damage promote recovery remains to be determined, but recent animal studies have shown that temporary silencing of retina ganglion cell activity in one or both eyes enables rapid recovery from deprivation amblyopia in adults that persists when FE activity is restored [16–19]. These observations are readily explained by the temporary lowering of the modification threshold for plasticity [20]. Accordingly, FE optic neuropathy (ION) and retinal pathologies should therefore serve as relatively strong drivers of AE recovery, even in adulthood.

Our findings also suggest that VA may not capture the full extent of AE improvement following FE injury. While VA is typically used to characterize the severity of amblyopia and response to treatment [21], multiple other visual deficits define the amblyopic state [22, 23], including decreased perimetric sensitivity [24]. All patients with available and reliable visual field data demonstrated clinically meaningful improvement in AE perimetric sensitivity. The magnitude and consistency of improvement in AE mean sensitivity argue against significant artifactual contributions from test-retest variance and/or familiarity with HVF testing [25, 26]. While our analysis was limited by sample size, it is clear that there is no relationship between improvements in perimetric performance and VA, suggesting central and peripheral AE visual gains may be independent in the setting of FE ION.

Unlike previous, largely isolated reports of AE improvement with FE pathology, our study design allows us to estimate the rate of AE recovery following FE ION. To our knowledge, this is the first report of peripheral amblyopic visual recovery following FE injury, thereby expanding the clinical phenotype of amblyopia recovery in adulthood. Limitations of this study are largely inherent to the nature of this phenomenon. The rarity of this condition limited the power with which we could assess hypothesized clinical relationships. Despite this, unambiguous, clinically meaningful trends emerged from our analysis. The retrospective design, albeit the only feasible mode to study this phenomenon, introduces inconsistencies in visual functional measurements, follow-up intervals, and reliability of clinical detail that may introduce bias. To specifically mitigate the concern for sampling bias with multiple VA measurements, we used the best reported AEVA in the 5 years preceding contralateral ION where possible to provide conservative measures of improvement. Additionally, our study examined patients receiving care at a tertiary ophthalmology center and this group may not be representative of the population at large; our identification of cases via database query and our inclusion of patients regardless of outcome, however, make this study more generalizable than any prior report of amblyopia following contralateral ION. Another major concern was potential confounding by change in AE refractive correction, especially since 7/8 cases with documented refractive correction who demonstrated AEVA improvement received an interval update. Improvement in AE function may have enabled these patients to provide more accurate responses during manifest refractions, and/or providers may have been more motivated to provide accurate AE correction following FE visual loss. Among studies of AE recovery following FE injury, this is among the few to examine change in refractive correction over time [6]. Nevertheless, the small magnitude of change in AE refractive correction, and near universal employment of pinhole VA argue against a significant refractive contribution to the AEVA gains observed in our cohort.

In conclusion, partial recovery of AE visual function is common and clinically significant in the setting of FE ION. Further study is needed to integrate these findings with prior reports of AE recovery, from which we may glean some important pathophysiologic insights into the neuroscientific bases for adult plasticity as it relates to visual recovery.

## Data Availability

All data produced in the present work are contained in the manuscript

## Figure Legends

**Supplemental Figure 1. Baseline AEVA and FEVA nadir do not share relationships with AEVA improvement**. (**A**) Change in AEVA after FE ION plotted as a function of baseline AEVA for all 12 cases. (**B**) Change in AEVA after FE ION plotted as a function of FEVA nadir for all 12 cases. LogMAR = logarithm of minimum angle of resolution.

**Supplemental Figure 2. Pattern of change in AE focal perimetric sensitivity does not match FE defects**. Map of AE HVF sensitivity change (dB) (left) with corresponding FE HVF gray scale image from first available HVF following FE ION onset (right) for the 6/12 cases with reliable AE HVF data. Change at each point was calculated using the raw sensitivity difference between the first and the most recent, reliable HVF.

