## Supplementary figures and images for "Partial recovery of amblyopia following fellow eye ischemic optic neuropathy"

### Supplemental Figure 1

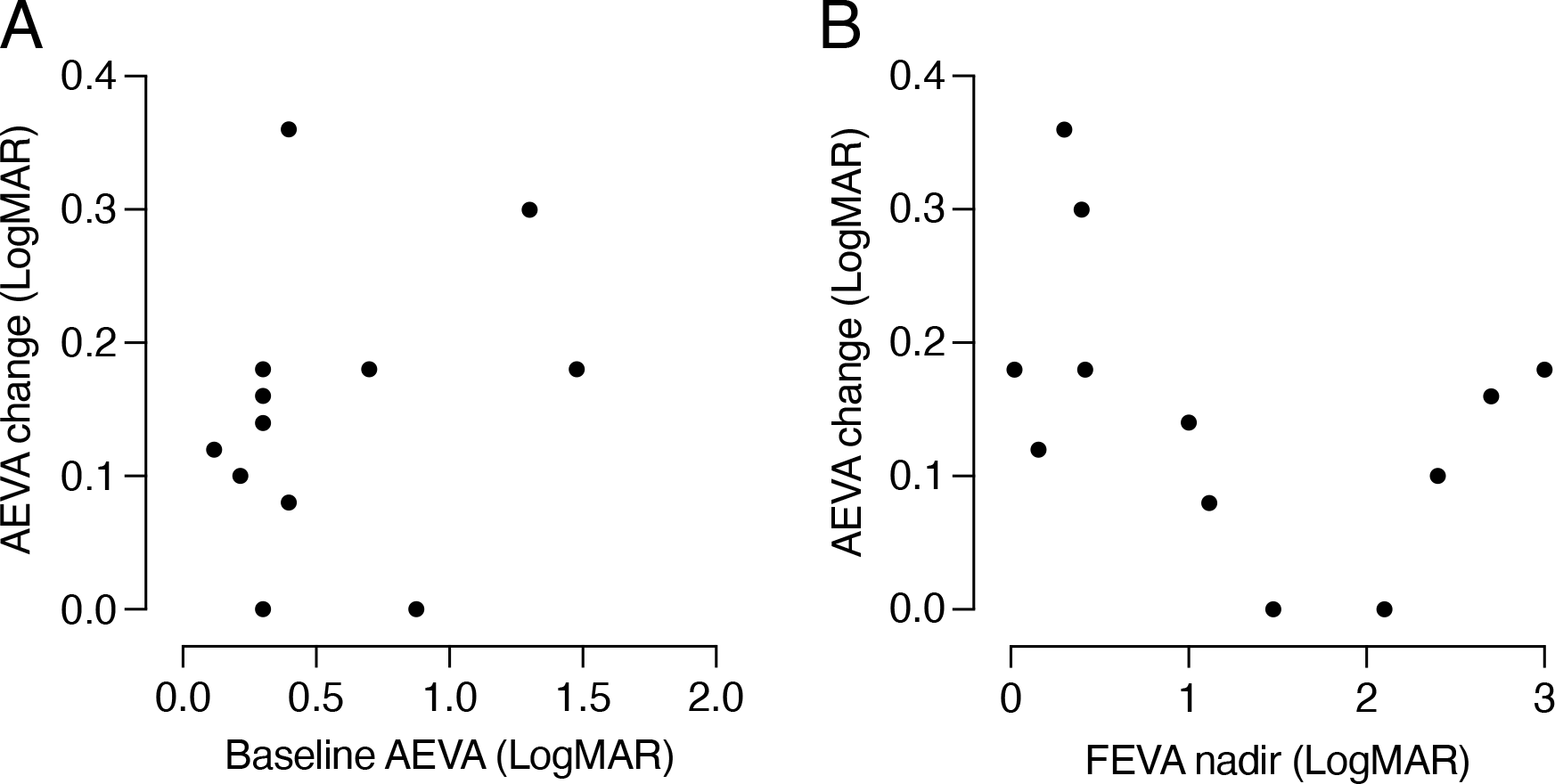

### Supplemental Figure 2

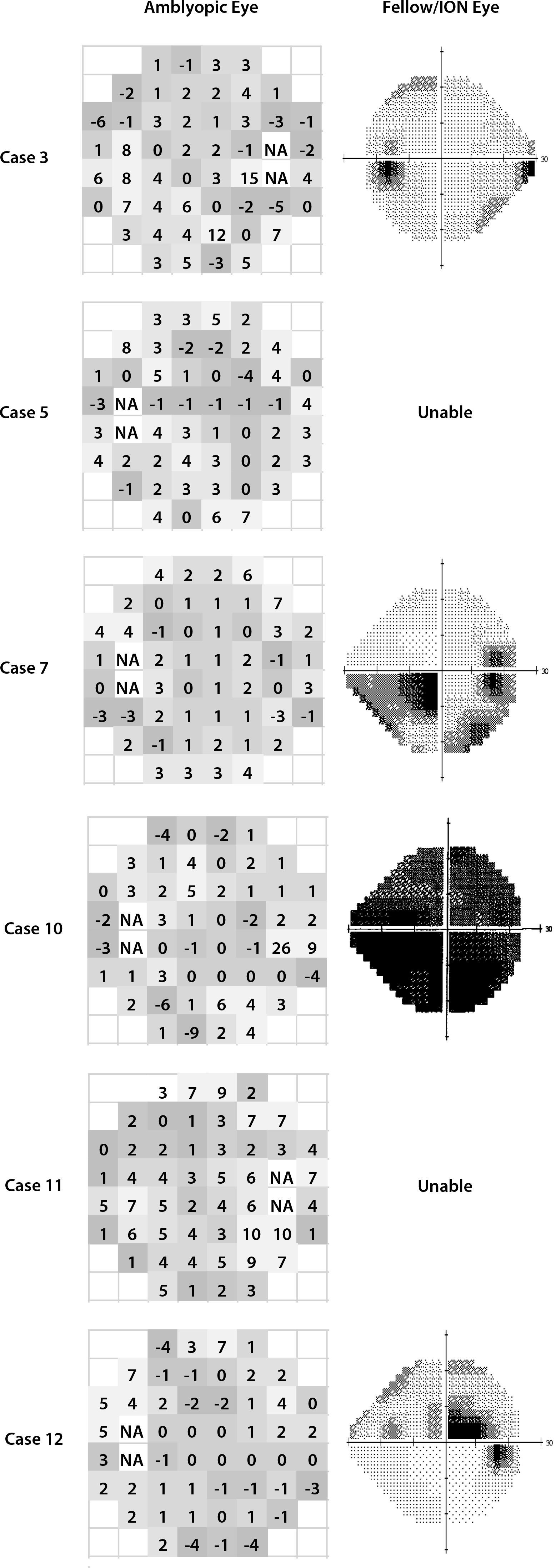
