## Supplemental Table 1 for "Partial recovery of amblyopia following fellow eye ischemic optic neuropathy"

| **Case #** | **Baseline Rx** | **Best AEVA Rx** | **Difference in spherical equivalent** |
| --- | --- | --- | --- |
| 1 | +7.00-2.50x79 | +8.75+2.00x180 | 4 |
| 3^a^ | sc PH | +1.25-0.75x75 | NA |
| 4 | +7.00-0.75x35 | +7.00-0.75x35 | 0 |
| 5 | +1.25 sph | sc PH | NA |
| 7 | +4.00-1.75x85 | +3.75-1.75x85 | -0.25 |
| 9 | +2.50 (balance lens) | PH^b^ | NA |
| 10 | sc (PH NI) | sc PH | NA |
| 11 | +3.00^c^ | +5.75-0.25x175 | 2.625 |
| 12 | Unknown | Unknown | Unknown |

**Supplemental Table 1.** Changes in AE refractive correction from baseline AEVA to best AEVA for those with ≥1 line of AE improvement

^a^ History of LASIK surgery with mono-vision (AE near, FE far)

^b^ Documentation unclear if VA measured with correction or with PH only; MRx at this visit is +1.75 sphere, but VA with MRx is unknown

^c^ Documented as using +3.00 over-the-counter readers; MRx measured at this visit as+2.50+2.25x85, but and produces worse VA than over-the-counter prescription

AE= amblyopic eye visual acuity; VA = Visual acuity; sc = sans correction; PH = pinhole; NI= no improvement
